# The causal role of thyroid hormones in bipolar disorders: a two-sample Mendelian Randomization study

**DOI:** 10.1101/2024.09.09.24313152

**Authors:** James L. Li

## Abstract

**Introduction:** Bipolar disorder is a complex psychiatric condition with distinctions between clinical subtypes including Type 1 and 2 disorders. Several studies have proposed thyroid hormones may be involved in the etiology of bipolar disorders.

**Methods:** This study employed a two-sample Mendelian Randomization (MR) approach to investigate the causal relationships between six thyroid hormone metrics (TSH, FT4, FT3, TT3, FT3/FT4, and TT3/FT4) and bipolar disorder, and Type 1 and 2 disorders, separately. Genome-wide association (GWAS) data from the Thyroidomics Consortium (up to 271,040 individuals of European ancestry) were used for thyroid function metrics. Bipolar disorder GWAS data included 41,917 cases and 371,549 controls (25,060 Type 1 and 6,781 Type 2 cases). We applied inverse variance weighted (IVW) methods for primary MR analysis, with MR Egger, weighted median, and weighted mode for sensitivity. Additional tests assessed horizontal pleiotropy and heterogeneity.

**Results:** Higher FT4 levels showed a protective causal effect against bipolar disorder (OR: 0.92, 95% CI: 0.86-0.97, p=4.58×10^−3^) and a suggestive effect on Type 1 disorders (OR: 0.92, 95% CI: 0.86-0.99, p=3.21×10^−2^). Elevated FT3 (OR: 1.18, 95% CI: 1.03-1.35, p=1.55×10^−2^) and FT3/FT4 ratio (OR: 1.97, 95% CI: 1.02-3.82, p=4.46×10^−2^) had suggestive harmful effects on Type 1 disorders. Sensitivity analyses showed consistent effects, with no significant horizontal pleiotropy or heterogeneity.

**Conclusions:** These findings highlight the protective role of FT4 and the potentially harmful effect of elevated FT3 in Type 1 bipolar disorder, highlighting the need for further research on thyroid hormone levels as a potential treatment strategy for Type 1 bipolar disorder.

## INTRODUCTION

Bipolar disorder is a complex psychiatric condition characterized by mood fluctuations that include episodes of mania or hypomania and depression^1^. The disorder is classified into two main subtypes: Type 1 bipolar disorder, which is marked by severe manic episodes, and Type 2, which involves less intense hypomanic episodes alongside depressive episodes^1^. Although the exact etiology of bipolar disorder remains unclear, some observational studies have identified biomarkers, including levels of thyroid hormones, that are associated with the disorder^2,3^; however, because of possible confounding factors, observational studies cannot establish a definitive causal link between thyroid function and bipolar disorders.

Mendelian Randomization (MR) has emerged as powerful tool for assessing the causal effects of exposures on outcomes by utilizing genetic variants as instruments^4^. While previous MR studies have suggested an association between free thyroxine (FT4) and overall bipolar disorder status^5,6^, the relationship between other thyroid hormones and bipolar disorder subtypes remains unexplored, in part due to limited sample sizes used in earlier genome-wide association studies (GWAS) for thyroid functions. Furthermore, the direct causal relationship between thyroid hormones and specific subtypes of bipolar disorders have remained largely unexplored.

In this study, we conduct a two-sample MR analysis using recently published GWAS summary statistics^7,8^ involving individuals with European ancestry to assess the potential causal link between thyroid hormones (exposures) and bipolar disorders (outcomes). We specifically examine six thyroid hormone metrics: Thyroid-stimulating hormone (TSH), FT4, free triiodothyronine (FT3), total triiodothyronine (TT3), the ratio of FT3 to FT4 (FT3/FT4), and the ratio of TT3 to FT4 (TT3/FT4).

Furthermore, we extend our analysis to investigate the causal relationships between these thyroid hormones and Type 1 and 2 bipolar disorders.

## MATERIALS AND METHODS

### Obtaining GWAS summary statistics for thyroid hormones and subtypes of bipolar disorder

GWAS summary statistics for TSH, FT4, FT3, TT3, FT3/FT4, and TT3/FT4 were obtained from Sterenborg et al.^7^ and were based on 271,040, 119,120, 59,061, 15,829, 51,095, and 15,510 individuals with European ancestry, respectively, who were participants in the ThyroidOmics Consortium; these individuals were above 18 years of age, not using thyroid medications, and did not have any history of thyroid surgeries. In generating these summary statistics, TSH, FT4, FT3, and TT3 were treated as continuous variables following an inverse normal transformation, and FT3/FT4 and TT3/FT4 underwent natural logarithm transformation^7^. Summary statistics for bipolar disorder, as well as Type 1 and 2 bipolar disorders were obtained from separate studies detailed in Mullins et al.^8^ involving a total of 41,917 bipolar disorder cases and 371,549 controls with European ancestry; of these cases, 25,060 individuals diagnosed with Type 1 bipolar disorder and 6,781 individuals diagnosed with Type 2 bipolar disorder were included for generating subtype-specific summary statistics. Diagnoses of bipolar disorder in most of these cases were based on the international consensus criteria for lifetime bipolar disorder (DSM-IV, ICD-9, or ICD-10), which were determined through structured diagnostic interviews, clinician-administered checklists, or medical record reviews^8^. All original studies contributing to these GWAS obtained ethical approval, and participants gave informed consent.

### Study design of the two-sample Mendelian Randomization (MR)

For the causal estimates derived from a Mendelian Randomization (MR) study to be considered valid, three key assumptions must be fulfilled: (1) the genetic variants must be strongly correlated with the exposure, (2) the genetic variants should not be linked to any confounders of the exposure-outcome relationship, and (3) the variants must not influence the outcome independently of the exposure. In this study, we employed a two-sample MR approach utilizing GWAS summary statistics to assess the association between each thyroid hormone (exposures) on each bipolar subtype (outcomes), utilizing genetic variants as instrumental variables.

### Selection of genetic variants as instruments

Separately for each thyroid hormone, we identified genetic variants associated with a p-value<5×10^−8^ and performed linkage disequilibrium clumping using PLINK 1.9^9^with a stringent threshold of r²<0.001 and a 10,000 kb window; European individuals from the 1000 genomes project^10^ were utilized as the reference panel for clumping. For thyroid hormones for which we identified very few genetic variants after clumping, we loosened the p-value threshold for clumping to 5×10^−7^ and 5×10^−6^ to obtain at least 10 initial genetic variants per thyroid hormone. Any genetic variants that were associated with the outcome (p<0.05) were then excluded from MR analyses. To assess whether pleiotropic pathways confounded the MR results, we queried all instruments in the NHGRI-EBI Catalog^11^ to determine if any instruments were associated with potential confounders between thyroid function and bipolar disorders; variants which were associated with potential confounders (including psychiatric and neurological conditions, as well as properties related to neurobiology such as brain function) were excluded from our analyses. For the remaining variants, we calculated the F-statistics using the formula β²/SE² to assess the strength of the selected genetic instruments, ensuring that all instruments used in our MR analyses had sufficient strength (F>10). We determined the variance explained by a specific SNP using the formula 2β^2^×ƒ×(1-ƒ), where β represents the SNP effect on a given thyroid hormone metric and ƒ represents the effect allele frequency. We aligned the GWAS summary statistics for both exposure and outcome by matching them based on the effect alleles and excluded palindromic variants with ambiguous allele frequencies from the MR analyses^12,13^.

### Performing two-sample MR between thyroid hormone metrics and subtypes of bipolar disorder

The inverse-variance weighted (IVW) method with multiplicative random effects^14^ was then applied to combine SNP-specific causal estimates for each bipolar disorder using the TwoSampleMR v0.6.7^15^ package in R; the IVW method is comparable to a two-stage least squares or allele score analysis when using individual-level data, and is therefore regarded as a conventional approach for MR. Given the interrelatedness of thyroid hormone metrics, we utilized a threshold of p-value<8.33×10^−3^ from the IVW method to identifying causal associations after accounting for multiple testing, corresponding to an alpha of 0.05 divided by 6 tests for the six thyroid hormone metrics. We reported suggestive causal associations using a threshold of p-value<0.05.

### Sensitivity analyses and assessment of pleiotropy

In addition to IVW, we utilized the TwoSampleMR package to implement MR-Egger^16^, weighted median^17^, and weighted mode^18^ MR methods to assess directional consistency of MR-IVW estimates. To confirm the robustness of significant findings, we conducted the Cochran’s Q-statistic test for heterogeneity, as well as tests for non-zero MR-Egger intercepts^12^ and the MR-Pleiotropy RESidual Sum and Outlier (MR-PRESSO) global test to assess pleiotropy^19^. For both causal (p-value<8.33×10^−3^) and suggestive causal (p-value<0.05) relationships we identified, we evaluated the influence of individual or pleiotropic variants by both computing MR estimates separately for each instrument, as well as conducting a leave-one-out analysis, where each SNP was systematically removed one at a time^16^ as a sensitivity analysis. We also evaluated whether our study had sufficient power to detect the associations we observed between thyroid hormone metrics and bipolar disease by performing power calculations using the mRnd webtool^20^. Lastly, we assessed whether there was publication bias in the summary statistics utilized for determining MR instruments by creating funnel plots for the causal associations we identified.

## RESULTS

### Genetic variants selected as instruments for each thyroid hormone metric

We identified genetic variants for each thyroid hormone metric (FT3, FT4, TSH, TT3, FT3/FT4, and TT3/FT4) that met the assumptions to serve as instruments in MR analyses including a total of 52 variants for FT4, 152 for TSH, 12 for FT3/FT4, and less than 10 variants for either FT3, TT3, or TT3/FT4 at a p-value threshold of 5×10^−8^ (**Supplementary Table 1**). When relaxing the p-value threshold to 5×10^−7^, we identified 13 suitable instruments for FT3 and 11 for TT3/FT4, and further relaxing the threshold to 5×10^−6^ provided 11 variants that qualified as instruments for TT3 (**Supplementary Table 1**). In total, we identified 8 genetic variants among all instruments across the six thyroid hormone metrics that were associated with pleiotropic pathways in the NHGRI-EBI GWAS Catalog^11^ that may confound the MR results, including psychiatric and neurological conditions, as well as traits related to brain function/anatomy (**Supplementary Table 2**); these 8 variants are not included in the aforementioned numbers of instruments nor in any subsequent MR analyses. F-statistics for all valid instrumental variables were greater than 10, ranging from 20.84 to 2183.15, confirming the strength and reliability of these instruments for MR analyses (**Supplementary Table 1**). Additional characteristics for the instruments utilized in MR analyses are presented in Supplementary Table 1.

### Two-sample Mendelian Randomization demonstrates thyroid hormones causally impact the risk of bipolar disorder subtypes

Utilizing genetic instruments for thyroid hormone metrics, we separately performed two-sample MR for overall bipolar disorder, as well as type 1 and type 2 bipolar disorders (**Supplementary Table 4**). We identified a causal negative relationship between FT4 hormone levels and the odds of bipolar disorder, overall, using the IVW method (OR: 0.92, 95% CI: 0.86, 0.97, p-value: 4.58×10^−3^) even after adjusting for multiple testing (**Table 1**). The directionality of this relationship between FT4 hormones and bipolar disorder identified by IVW was highly consistent across MR-Egger, weighted median, and weighted mode MR approaches (**Figure 1**). We also identified FT4 hormone levels had a suggestive causal negative relationship with Type 1 bipolar disorder (OR: 0.92, 95% CI: 0.86, 0.99, p-value: 3.21×10^−2^), which was also directionally consistent across other MR approaches (**Table 1 and Figure 1**). However, the MR association between FT4 and Type 2 bipolar disorder was not statistically significant (IVW p-value: 0.29), and directionality of this relationship differed between MR approaches (**Supplementary Table 4**). Furthermore, we identified two other thyroid hormone metrics, FT3 and FT3/FT4, had suggestive causal associations with Type 1 bipolar disorder yielding IVW ORs of 1.18 (95% CI: 1.03, 1.35, p-value: 1.55×10^−2^) and 1.97 (95% CI: 1.02, 3.82, p-value: 4.46×10^−2^), respectively (**Table 1**); these associations were directionally consistent across MR methods (**Table 1 and Figure 1**). While the associations between FT3/FT4 and Type 2 bipolar disorder were not statistically significant (p>0.05), the directionality of the association seemed to be opposite to that between FT3/FT4 and Type 1 bipolar disorder (**Supplementary Table 4**). MR associations between FT3 and Type 2 bipolar disorder also appeared to be opposite in directionality compared to the association with Type 1 bipolar disorder, but these models showed evidence of directional pleiotropy (MR-Egger intercept p-value: 0.048).

**Table 1.** Causal and suggestive associations identified between thyroid hormone metrics and bipolar disorder subtypes in two-sample Mendelian Randomization analyses.

| Subtype | Thyroid Hormone | MR Method | Number of Instruments | Odds Ratio (95% CI) | P-value | Cochran's Q P-value | MR-egger Intercept P-value | MR-PRESSO Global Test P-value |
| --- | --- | --- | --- | --- | --- | --- | --- | --- |
| Bipolar Disorder (overall) | FT4 | IVW | 43 | 0.92<br>(0.86, 0.97) | 4.58E-03* | 0.81 | 0.69 | 0.83 |
|  |  | MR Egger | 43 | 0.89<br>(0.78, 1.02) | 0.11 |  |  |  |
|  |  | Weighted median | 43 | 0.89<br>(0.80, 0.98) | 2.43E-02 |  |  |  |
|  |  | Weighted mode | 43 | 0.87<br>(0.79, 0.97) | 1.30E-02 |  |  |  |
| Type 1 Bipolar Disorder | FT3 | IVW | 11 | 1.18<br>(1.03, 1.35) | 1.55E-02** | 0.79 | 0.75 | 0.85 |
|  |  | MR Egger | 11 | 1.09<br>(0.68, 1.75) | 0.72 |  |  |  |
|  |  | Weighted median | 11 | 1.15<br>(0.93, 1.43) | 0.19 |  |  |  |
|  |  | Weighted mode | 11 | 1.14<br>(0.90, 1.45) | 0.30 |  |  |  |
|  | FT4 | IVW | 36 | 0.92<br>(0.86, 0.99) | 3.21E-02** | 0.95 | 0.80 | 0.96 |
|  |  | MR Egger | 36 | 0.94<br>(0.79, 1.12) | 0.49 |  |  |  |
|  |  | Weighted median | 36 | 0.90<br>(0.79, 1.04) | 0.15 |  |  |  |
|  |  | Weighted mode | 36 | 0.90<br>(0.78, 1.03) | 0.13 |  |  |  |
|  | FT3/FT4 | IVW | 7 | 1.97<br>(1.02, 3.82) | 4.46E-02** | 0.32 | 0.41 | 0.46 |
|  |  | MR Egger | 7 | 1.24<br>(0.37, 4.18) | 0.74 |  |  |  |
|  |  | Weighted median | 7 | 1.61<br>(0.79, 3.28) | 0.19 |  |  |  |
|  |  | Weighted mode | 7 | 1.62<br>(0.80, 3.27) | 0.23 |  |  |  |

*Note:* Odds ratios are based on FT4 and FT3 levels following an inverse normal transformation and FT3/FT4 undergoing a natural logarithm transformation.
\* Causal association identified at multiple testing threshold (Bonferroni) of $p\text{-value} < 8.33 \times 10^{-3}$
\*\* Suggestive causal association identified at a nominal significance threshold of $p\text{-value} < 0.05$

**Figure 1.**
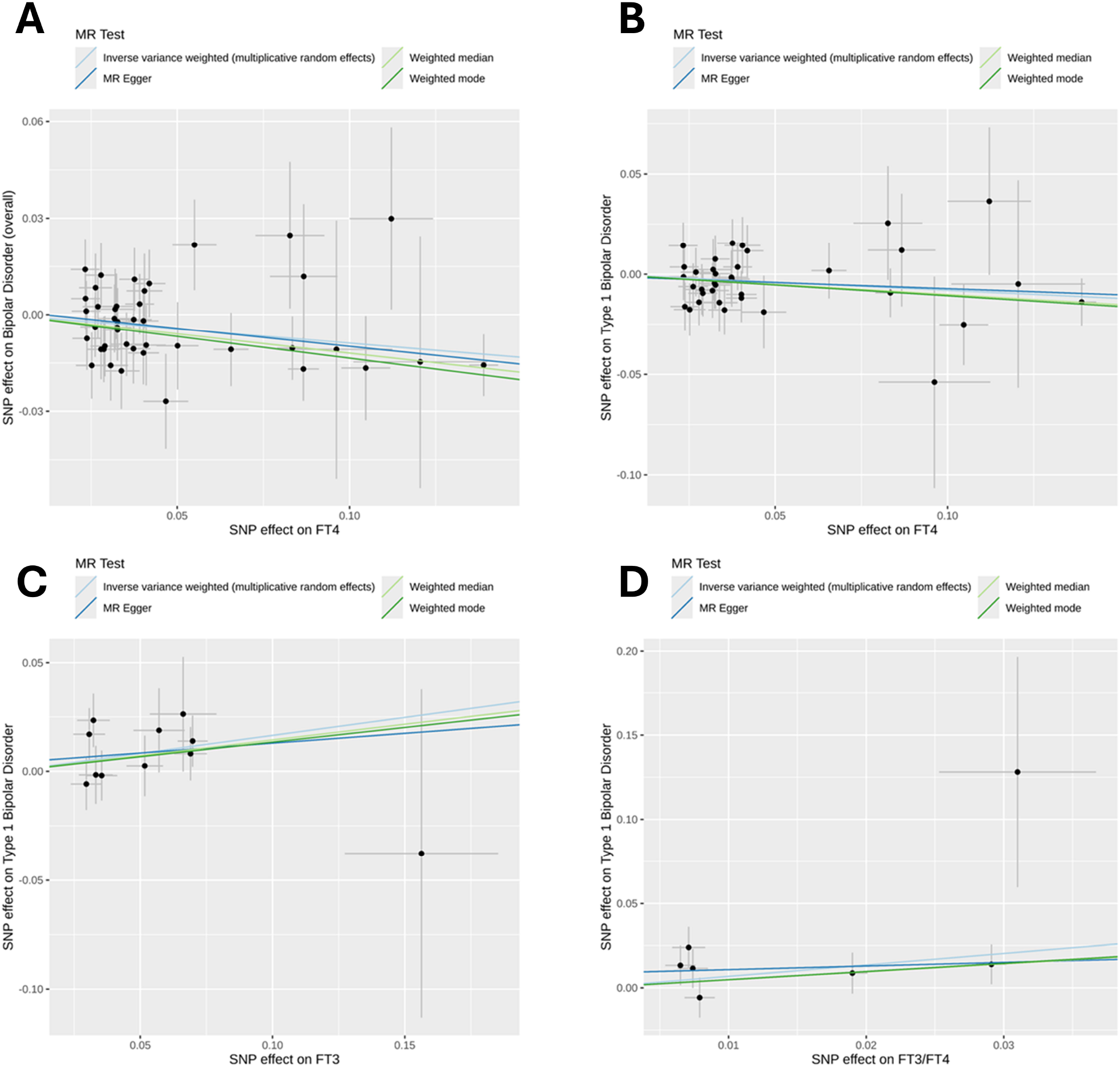
Scatterplots for the effects of instruments on each thyroid hormone metric and bipolar disorder subtype for causal and suggestive associations identified in Mendelian Randomization (MR) analyses. A) Relationship between FT4 and bipolar disorder (overall), B) Relationship between FT4 and Type 1 bipolar disorder, C) Relationship between FT3 and Type 1 bipolar disorder, D) Relationship between FT3/FT4 and Type 1 bipolar disorder.

For the one causal and three suggestive causal relationships we identified between thyroid hormone metrics and bipolar disorder subtypes (**Table 1**), we rigorously assessed potential biases by conducting several sensitivity analyses. The instruments utilized in these relationships did not show evidence of being in any confounding or pleiotropic pathways when examining genome-wide significant associations between these instruments and diseases/traits cataloged in NHGRI-EBI GWAS Catalog (**Supplementary Table 4**). We did not find evidence for pleiotropy across instrument effects (Cochran’s Q test p-values>0.05) (**Table 1**). Next, there was no evidence of directional pleiotropy among these causal associations (MR-Egger intercept p-values>0.05) (**Table 1**). In addition, we computed the MR associations for each instrument separately and observed that they were generally consistent with the estimates inferred by each MR method collectively across all instruments (**Figure 2**). Moreover, MR-PRESSO did not detect any outlier genetic variants that needed to be removed (MR-PRESSO global test p-values>0.05) (**Table 1**). To further corroborate the absence of outlier instruments, we performed a leave-one-out analysis where we computed MR estimates but left each instrument one-by-one; we observed our findings were generally reliable and stable as none of the instruments heavily influenced the causal relationships we observed when excluded (**Figure 3**).

**Figure 2.**
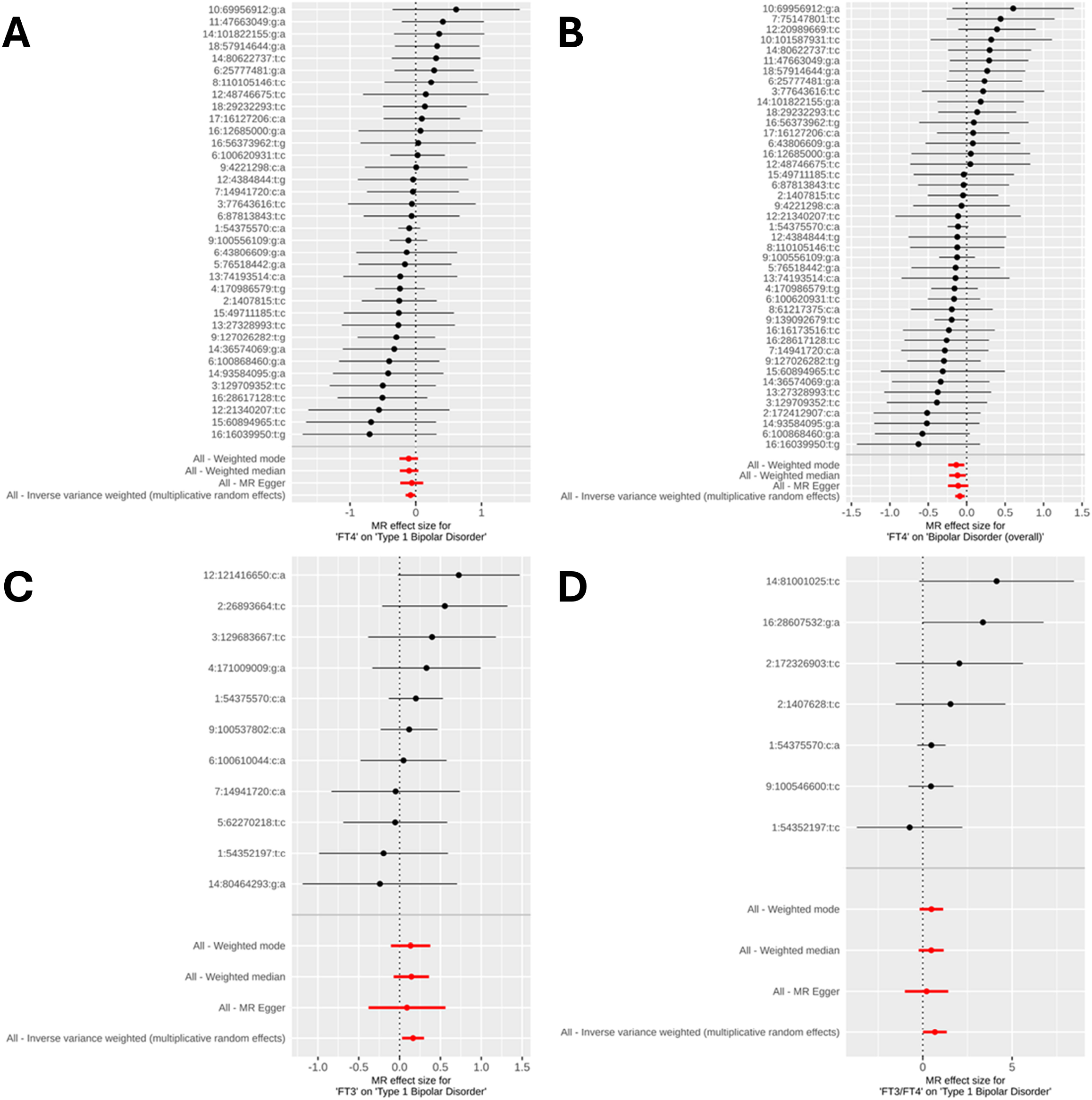
Forest plots of the Mendelian Randomization (MR) estimates for each instrument individually compared to MR approaches using all instruments collectively. A) Relationship between FT4 and bipolar disorder (overall), B) Relationship between FT4 and Type 1 bipolar disorder, C) Relationship between FT3 and Type 1 bipolar disorder, D) Relationship between FT3/FT4 and Type 1 bipolar disorder.

**Figure 3.**
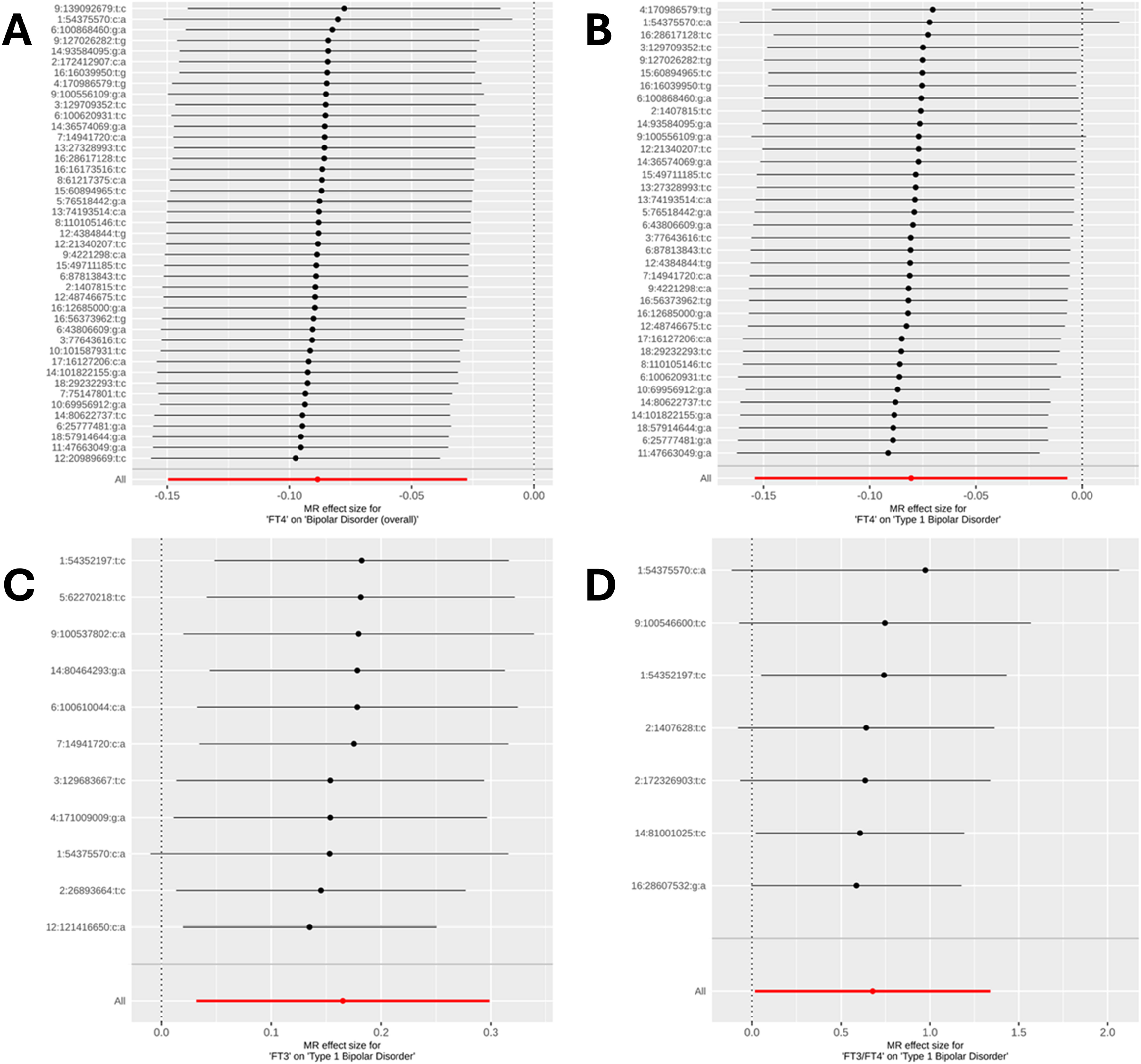
Forest plots of the inverse-variance weighted Mendelian Randomization (MR) method estimates utilizing the leave-one-out approach where instruments were excluded one-by-one. A) Relationship between FT4 and bipolar disorder (overall), B) Relationship between FT4 and Type 1 bipolar disorder, C) Relationship between FT3 and Type 1 bipolar disorder, D) Relationship between FT3/FT4 and Type 1 bipolar disorder.

Additionally, we conducted power calculations for sample sizes of 413,466 (41,917 cases and 371,549 controls) for bipolar disorder overall and 396,609 (25,060 cases and 371,549 controls) for Type 1 bipolar disorder, with an alpha of 0.05. The sum of variance in FT4 explained by instruments in the MR with bipolar disorder overall was 3.84%, yielding a power of 0.87 to detect the MR association we observed. For Type 1 bipolar disorder, the sum of variance in FT4, FT3, and FT3/FT4 explained by instruments was 3.20%, 0.96%, and 0.07%, respectively, yielding power values of 0.60, 0.76, and 0.97, respectively, for detecting the MR associations we observed. Funnel plots for each of the causal relationships we identified also suggested that there were no indications of publication bias in this study (**Supplementary Figure 1**).

## DISCUSSION

In this study, we conducted a two-sample MR of six thyroid hormone metrics and bipolar disorder overall, as well as Type 1 and 2 bipolar disorders. This study suggests that FT4 hormone levels may have a protective causal effect on bipolar disorder overall, with a suggestive causal effect on Type 1 bipolar disorder. However, we also identified suggestive harmful causal effects of elevated FT3 levels and a higher FT3/FT4 ratio on Type 1 bipolar disorder. These findings remained consistent across various MR methods, each with different assumptions about horizontal and directional pleiotropy, suggesting that pleiotropy is unlikely to account for our results.

Our study aligns with previous Mendelian randomization research showing that FT4 has a protective impact on the risk of bipolar disorder^5,6^ and supports observational studies that found lower FT4 levels in bipolar disorder patients compared to controls^21^; however, our research specifically suggests this relationship may be with Type 1 bipolar disorder. Although we cannot entirely dismiss the possibility that FT4 influences Type 2 bipolar disorder, this uncertainty is largely due to the limited sample size of the Type 2 bipolar disorder GWAS summary statistics^8^ used in our MR analysis. Additionally, our study reveals novel suggestive causal associations, showing a negative relationship between Type 1 bipolar disorder and both FT3 and the FT3/FT4 ratio. Notably, the opposing directionalities of the association between FT4 and Type 1 bipolar disorder and that between FT3, FT3/FT4, and Type 1 bipolar disorder are consistent with the conversion process of FT3 to FT4 in the body. This finding suggests that the conversion of FT3 to FT4 may in part have a protective effect against bipolar disorder; however, further research is needed to determine whether this protective effect is solely due to increased FT4 production or if it is also influenced independently by decreased FT3 levels.

Furthermore, by validating previously reported MR associations between FT4 and bipolar disorder using much larger GWAS summary statistics for FT4^7^, with an approximately 65% increase in the number of participants compared to the latest prior study^5^, our study provides more confidence that FT4 supplementation should be considered in the treatment of patients with Type 1 bipolar disorder. Interestingly, a recent clinical trial that administered levothyroxine and triiodothyronine as adjunctive treatments for patients who did not respond to lithium therapy demonstrated that levothyroxine had a positive effect on reducing mixed states^22^. Given that our results suggest a reduction in FT3 levels may decrease the odds of Type 1 bipolar disorder, follow-up studies designed to explicitly evaluate the role of lowering FT3 while increasing FT4 levels are warranted.

Though this study managed to identify causal and suggestive causal relationships between thyroid functioning and bipolar disorders, there are several limitations. First, the sample size of individuals used to generate GWAS summary statistics for Type 2 bipolar disorder were quite underpowered^8^ and may have potentially contributed to the lack of associations between thyroid hormone metrics and this subtype. Additionally, thyroid hormone measurements are quite variable even when taken from the same individual at different time points, including during the daytime compared to in the evening^23^. Restricting participants included in the generation of thyroid function GWAS summary statistics to only those in a subset of studies with identical sample collection protocols (such as standardized time points for measurements) may help increase the precision of MR estimates. Furthermore, this study was constrained to individuals with European ancestry since GWAS summary statistics for both thyroid function metrics and bipolar disorder are quite limited in non-European populations; validation studies in other non-European populations are warranted.

Overall, these results contribute to a deeper understanding of the endocrine factors involved in bipolar disorders and underscore how both FT4 and FT3 may impact the risk of Type 1 bipolar disorder. These findings further suggest that thyroid hormone metrics could potentially serve as biomarkers or therapeutic targets in the management of bipolar disorders.

## Data Availability

Code for all analyses performed in this study are available at https://github.com/james-li-projects/MR_Thyroid_BPD.

## AUTHOR CONTRIBUTIONS

J.L.L. designed the study, performed data processing, performed data analysis, and wrote the manuscript.

## ACKNOWLEDGEMENTS

The author would like to thank the support staff of Randi, a high-performance computing cluster maintained by the University of Chicago’s Center for Research Informatics.

## TRANSPARENCY DECLARATION

The author affirms the manuscript provides a truthful, accurate, and transparent account of the reported study, with no significant aspects omitted.

## DATA AND CODE AVAILABILITY STATEMENT

All GWAS summary statistics utilized in this study are previously published and publicly available and for academic use without restriction. GWAS summary statistics for thyroid function utilized in this study are downloadable from the Thyroidomics Consortium at https://transfer.sysepi.medizin.uni-greifswald.de/thyroidomics/datasets/. GWAS summary statistics for bipolar disorders utilized in this study are downloadable from the Psychiatric Genomics Consortium at https://pgc.unc.edu/for-researchers/download-results/. Code for all analyses performed in this study are available at https://github.com/james-li-projects/MR_Thyroid_BPD.

## Supplementary Tables and Figures

**Supplementary Figure 1.** Funnel plots for the instruments utilized in each of the four causal relationships between thyroid hormones and bipolar disorder subtypes we identified. A) Relationship between FT4 and bipolar disorder (overall), B) Relationship between FT4 and Type 1 bipolar disorder, C) Relationship between FT3 and Type 1 bipolar disorder, D) Relationship between FT3/FT4 and Type 1 bipolar disorder.

**Supplementary Table 1.** Characteristics of genetic variants used as instruments in the Mendelian Randomization between each thyroid hormone metric and bipolar disorder subtype.

**Supplementary Table 2.** Previously reported GWAS associations for the 8 invalid instruments that were excluded due to their links with potential confounding traits or diseases.

**Supplementary Table 3.** Mendelian Randomization estimates between each thyroid hormone metric and bipolar disease subtype.

**Supplementary Table 4.** Previously reported GWAS associations for the valid instruments used in the identified causal and suggestive associations showed no evidence of links to potential confounding traits or diseases.

## REFERENCES

1. Grande I, Berk M, Birmaher B, Vieta E. Bipolar disorder. The Lancet. 2016;387(10027):1561–1572. doi:10.1016/S0140-6736(15)00241-X

2. Song X, Feng Y, Yi L, Zhong B, Li Y. Changes in thyroid function levels in female patients with first-episode bipolar disorder. Front Psychiatry. 2023;14. doi:10.3389/fpsyt.2023.1185943

3. Chakrabarti S. Thyroid Functions and Bipolar Affective Disorder. J Thyroid Res. 2011;2011:306367. doi:10.4061/2011/306367

4. Lawlor DA, Harbord RM, Sterne JAC, Timpson N, Davey Smith G. Mendelian randomization: using genes as instruments for making causal inferences in epidemiology. Stat Med. 2008;27(8):1133–1163. doi:10.1002/sim.3034

5. Chen G, Lv H, Zhang X, et al. Assessment of the relationships between genetic determinants of thyroid functions and bipolar disorder: A mendelian randomization study. J Affect Disord. 2022;298:373–380. doi:10.1016/j.jad.2021.10.101

6. Gan Z, Wu X, Chen Z, et al. Rapid cycling bipolar disorder is associated with antithyroid antibodies, instead of thyroid dysfunction. BMC Psychiatry. 2019;19(1):378. doi:10.1186/s12888-019-2354-6

7. Sterenborg RBTM, Steinbrenner I, Li Y, et al. Multi-trait analysis characterizes the genetics of thyroid function and identifies causal associations with clinical implications. Nat Commun. 2024;15(1):888. doi:10.1038/s41467-024-44701-9

8. Mullins N, Forstner AJ, O’Connell KS, et al. Genome-wide association study of more than 40,000 bipolar disorder cases provides new insights into the underlying biology. Nat Genet. 2021;53(6):817–829. doi:10.1038/s41588-021-00857-4

9. Purcell S, Neale B, Todd-Brown K, et al. PLINK: A Tool Set for Whole-Genome Association and Population-Based Linkage Analyses. Am J Hum Genet. 2007;81(3):559–575.

10. Auton A, Abecasis GR, Altshuler DM, et al. A global reference for human genetic variation. Nature. 2015;526(7571):68–74. doi:10.1038/nature15393

11. Sollis E, Mosaku A, Abid A, et al. The NHGRI-EBI GWAS Catalog: knowledgebase and deposition resource. Nucleic Acids Res. 2023;51(D1):D977-D985. doi:10.1093/nar/gkac1010

12. Bowden J, Davey Smith G, Burgess S. Mendelian randomization with invalid instruments: effect estimation and bias detection through Egger regression. Int J Epidemiol. 2015;44(2):512–525. doi:10.1093/ije/dyv080

13. Bowden J, Del Greco M F, Minelli C, Davey Smith G, Sheehan N, Thompson J. A framework for the investigation of pleiotropy in two-sample summary data Mendelian randomization. Stat Med. 2017;36(11):1783–1802. doi:10.1002/sim.7221

14. Burgess S, Butterworth A, Thompson SG. Mendelian randomization analysis with multiple genetic variants using summarized data. Genet Epidemiol. 2013;37(7):658–665. doi:10.1002/gepi.21758

15. Hemani G, Zheng J, Elsworth B, et al. The MR-Base platform supports systematic causal inference across the human phenome. Loos R, ed. eLife. 2018;7:e34408. doi:10.7554/eLife.34408

16. Burgess S, Thompson SG. Interpreting findings from Mendelian randomization using the MR-Egger method. Eur J Epidemiol. 2017;32(5):377–389. doi:10.1007/s10654-017-0255-x

17. Bowden J, Davey Smith G, Haycock PC, Burgess S. Consistent Estimation in Mendelian Randomization with Some Invalid Instruments Using a Weighted Median Estimator. Genet Epidemiol. 2016;40(4):304–314. doi:10.1002/gepi.21965

18. Hartwig FP, Davey Smith G, Bowden J. Robust inference in summary data Mendelian randomization via the zero modal pleiotropy assumption. Int J Epidemiol. 2017;46(6):1985–1998. doi:10.1093/ije/dyx102

19. Verbanck M, Chen CY, Neale B, Do R. Detection of widespread horizontal pleiotropy in causal relationships inferred from Mendelian randomization between complex traits and diseases. Nat Genet. 2018;50(5):693–698. doi:10.1038/s41588-018-0099-7

20. Brion MJA, Shakhbazov K, Visscher PM. Calculating statistical power in Mendelian randomization studies. Int J Epidemiol. 2013;42(5):1497–1501. doi:10.1093/ije/dyt179

21. Vedal TSJ, Steen NE, Birkeland KI, et al. Free thyroxine and thyroid-stimulating hormone in severe mental disorders: A naturalistic study with focus on antipsychotic medication. J Psychiatr Res. 2018;106:74–81. doi:10.1016/j.jpsychires.2018.09.014

22. Walshaw PD, Gyulai L, Bauer M, et al. Adjunctive thyroid hormone treatment in rapid cycling bipolar disorder: A double-blind placebo-controlled trial of levothyroxine (L-T4) and triiodothyronine (T3). Bipolar Disord. 2018;20(7):594–603. doi:10.1111/bdi.12657

23. Mahadevan S, Sadacharan D, Kannan S, Suryanarayanan A. Does Time of Sampling or Food Intake Alter Thyroid Function Test? Indian J Endocrinol Metab. 2017;21(3):369–372. doi:10.4103/ijem.IJEM_15_17

