## Supplementary figures and images for "The causal role of thyroid hormones in bipolar disorders: a two-sample Mendelian Randomization study"

### Supplementary Figure 1

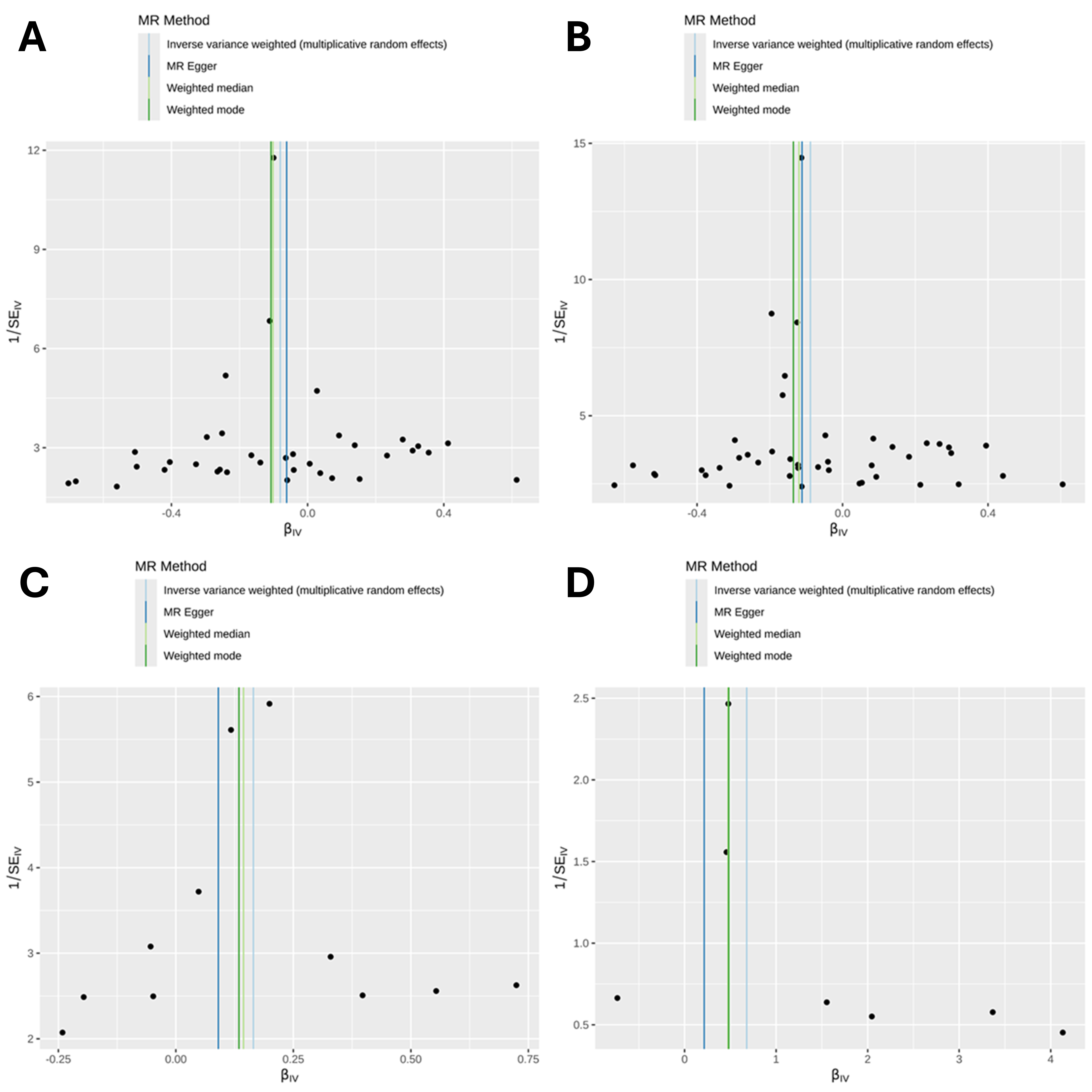
